# Mechanically assisted walking training during inpatient rehabilitation after stroke: perceptions of physiotherapists

**DOI:** 10.1101/2020.12.10.20247288

**Authors:** Sandra G Brauer, Lauren Waters, Suzanne S Kuys, Louise Ada

## Abstract

**Background and Purpose:** Despite evidence supporting the use of mechanically assisted walking training in stroke rehabilitation, it is not extensively used. The purpose of this study was to explore the perceptions of physiotherapists regarding their use of mechanically assisted walking training, specifically treadmill training, during inpatient rehabilitation after stroke. Better understanding of physiotherapists’ perceptions can inform the development of translation strategies.

**Methods:** A qualitative study using focus groups comprising 14 physiotherapists (including students) working in stroke inpatient rehabilitation at two sites was conducted. Transcripts were analysed using an inductive approach to thematic analysis.

**Results:** Physiotherapists perceived two main themes related to using mechanically assisted walking training during inpatient rehabilitation: therapeutic consequences (eg, patients able to walk earlier, further, longer; less fatiguing for therapist, ability to manipulate walking parameters) and practical considerations (eg, safety, efficiency, staff skill, access to equipment, weight and level of disability of patient, fear of treadmill).

**Discussion:** Therapists’ perceptions of using mechanically assisted walking training during inpatient rehabilitation after stroke were mixed. There is a need to educate physiotherapy staff about the evidence of therapeutic value as well as how to perform mechanically assisted walking training.

## INTRODUCTION

Maximising walking ability is a priority in the acute and sub-acute phases of rehabilitation after stroke. At the time of this study, there was evidence that for ambulatory stroke survivors, mechanically assisted walking training without body weight support lead to clinically meaningful improvements in walking speed and distance (Polese, Ada, Dean, Nascimento, & Teixeira-Salmela, 2013) without adversely affecting walking quality (Kuys, Brauer, & Ada, 2011). There was also evidence that, non-ambulatory stroke survivors who received mechanically assisted walking training with body weight support during rehabilitation were more likely to return to independent walking than those who received traditional overground gait retraining (Ada, Dean, Vargas, & Ennis, 2010). Therefore, the Australian clinical guidelines for stroke management at the time in 2010 (National Stroke Foundation, 2010) recommended that:

> *People with difficulty walking should be given the opportunity to undertake tailored, repetitive practice of walking as much as possible and that mechanically assisted gait* (*via treadmill or automated mechanical or robotic device*) *be used in addition to conventional walking training*.

Despite the evidence supporting its use, and recommendations for its uptake in clinical practice guidelines, there was little indication that mechanically assisted walking training was extensively used in stroke rehabilitation. In an Australian national stroke audit of rehabilitation services in 2018, only 17% of people unable to walk independently received mechanically assisted gait training of any type (Stroke Foundation, 2018) up from 14% in 2016 (Stroke Foundation, 2016). Effecting knowledge transfer is complex, with adoption of innovation influenced by the innovation itself, the recipients and the context (Graham & Logan, 2004). We chose to examine therapists’ perceptions regarding the use of mechanically assisted walking training during inpatient rehabilitation for stroke survivors. The mechanically assisted walking devices available within the settings involved in this study were treadmill and body weight support systems. Therefore, the aim was to gain insight into facilitators and barriers to mechanically assisted walking training, specifically treadmill and body weight support, to inform strategies to optimise translation into clinical practice in the future. This study addresses the need for more understanding about the translation of evidence into practice in stroke rehabilitation (Lynch, Chesworth, & Connell, 2018).

## METHODS

### Design

A descriptive qualitative design was used. Focus groups were undertaken with physiotherapists working in inpatient rehabilitation to explore their perceptions of using mechanically assisted walking training to improve walking after stroke. Focus groups were employed to collect data as they encourage and promote discussion between colleagues and can provide insights that individual interviews may miss. Additionally, focus groups are commonly used to explore implementation of new technologies (Davidson, Halcomb, & Gholizadeh, 2017). Hospital and university human medical research ethics committees granted approval to conduct the research. All participants provided written informed consent. The study was conducted in accordance with the Declaration of Helsinki (World Medical Association, 2013).

### Setting

Focus groups were conducted at two rehabilitation sites located in Queensland, Australia. Site A was a 30-bed general rehabilitation unit situated within a tertiary hospital. Site B was a 51-bed independent rehabilitation unit. At both sites, physiotherapy services were provided during weekdays only, by about 10 staff: a core group of therapists on a permanent basis, therapists on three-month rotation, as well as students. For both sites mechanically assisted devices were a treadmill and body weight support system which was easily accessible in the therapy area. Both sites had a throughput of 150 stroke patients annually.

### Sampling and recruitment

All physiotherapists and physiotherapy students working in the rehabilitation units at each site were eligible to participate in the focus group to understand diverse experiences. At Site A, physiotherapists who had worked in the rehabilitation unit during their last rotation (but who were now working elsewhere in the hospital) were also asked to participate. Participants were approached via email after a face-to-face meeting between the investigators (SB and SK) and all physiotherapy staff currently working in the unit.

### Data collection

A single focus group lasting one hour was conducted at each site in a private room away from the rehabilitation gyms. The open-ended stimulus question was: *what are your perceptions about training stroke patients using the treadmill?* Probes explored perceptions for treadmill use with and without the harness and with and without body weight support. A facilitator, a physiotherapist with a PhD experienced in conducting focus groups who did not work at either site and had no association with any of the participants, conducted both focus groups (RL). The facilitator had previously worked as a senior physiotherapist in a similar inpatient rehabilitation setting but had no previous research with a stroke population and did not have strong viewpoints about the use of mechanically assisted walking training. The role of the facilitator was to ask probing questions in order to further explore pertinent topics, and to paraphrase dialogue to allow participants to concur with, add to, or dispute the main points raised. At the end of the discussion, the facilitator summarized the main issues that had been raised during the session and the participants were given the opportunity to make clarifications or contribute further information. A second researcher (LW) acted as a scribe, made comprehensive notes on group dynamics and non-verbal signals. The focus groups were voice recorded and transcribed verbatim. The facilitator read through the transcripts adding additional information or meaning gleaned through the role of facilitator.

### Data analysis

The inductive approach to thematic data analysis described by Braun and Clarke (2006) was used to identify recurring themes within the data. While we describe the process in phases, data analysis was not linear. We repeated tasks in each phase until agreement relating to the findings was reached. In phase one, two of the researchers involved in the study (LW and SB) familiarised themselves with the data by independently reading and re-reading the focus group transcripts. While LW was involved in the data collection process, SB was not. The intention of the first reading was not to generate codes but to gain an appreciation of the data in its entirety. In the second reading, the reviewers took notes about possible patterns and meanings apparent in the data. In phase two, the reviewers worked independently to ascribe codes to chunks of data that appeared to be meaningful or interesting. The reviewers then met to discuss similarities and differences between how they had coded the data. Codes sharing similar topics or meaning were grouped together into overarching themes (phase three). Data within each theme should be meaningful and coherent on its own, and there should also be a clear distinction between data in each theme (Braun & Clarke, 2006). Once a core set of themes had been established, the reviewers refined the themes (phase four). A third reviewer (SK), who did not take part in the data collection, was involved with this stage of the analysis. The purpose of this phase was to ensure that designated themes accurately reflected the data as a whole and that the codes within each theme formed a coherent pattern. If there was overlap between themes, themes were modified to more accurately represent the data. Participants provided feedback on the findings at a follow up meeting.

## RESULTS

A total of 14 participants were recruited: seven from Site A and seven from Site B, representing most of each site’s physiotherapy staff. Two people (one at each site) did not participate, due to being on leave on the day of the focus group. At each site, two physiotherapy students participated in the focus groups. Characteristics of the participants are presented in Table 1.

**Table 1.** Characteristics of participants

| Participant | Site | Position | Time since graduation (yr) | Clinical experience in rehabilitation (yr) |
| --- | --- | --- | --- | --- |
| 1 | A | Physiotherapist (senior) | 18 | 16 |
| 2 | A | Physiotherapist | 1.5 | 0.5 |
| 3 | A | Physiotherapist | 9.5 | 8 |
| 4 | A | Physiotherapist | 4.5 | 1.5 |
| 5 | A | Physiotherapist | 6.5 | 4 |
| 6 | A | Student | 0 | 0 |
| 7 | A | Student | 0 | 0 |
| 8 | B | Physiotherapist | 25 | 16 |
| 9 | B | Physiotherapist | 2.5 | 0.5 |
| 10 | B | Physiotherapist | 12 | 8 |
| 11 | B | Student | 0 | 0 |
| 12 | B | Student | 0 | 0 |
| 13 | B | Physiotherapist | 3.5 | 1.5 |
| 14 | B | Physiotherapist (senior) | 22 | 19 |

The discussions highlighted possible explanations for why a physiotherapist might choose to use, or not use, mechanically assisted walking training during stroke rehabilitation. Two major themes emerged: therapeutic consequences and practical considerations. These themes were often discussed in terms of being both barriers to and/or facilitators of the use of the treadmill (Table 2).

**Table 2.** Summary of therapists’ perceptions of use of mechanically assisted walking training during inpatient rehabilitation after stroke.

| Themes | Facilitators | Barriers |
| --- | --- | --- |
| Therapeutic consequences | Walk earlier<br>Walk further/longer<br>Manipulate walking parameters | Abnormal muscle activation |
| Practical considerations | More efficient for staff<br>More safe for patients<br>Less fatiguing for staff | Less efficient for staff<br>Less safe for patients<br>Staff skill is required<br>Large weight of patient<br>High level of disability of patient<br>Fear of treadmill by patient<br>Access difficult |

### Therapeutic consequences

There was general agreement that the treadmill could be of benefit to some patients who have had a stroke. It was suggested that using a treadmill could enable stroke survivors to walk earlier, and further, than they would be able to without the treadmill:

> *Early walking* …(*is a benefit of using the treadmill*) (*Participant 5*)
>
> *I think you can get a patient to walk further on a treadmill than what you ever could over land*. (*Participant 3*)

Participants also described how treadmill use can allow them to manually facilitate the patient’s walking for long periods of time without becoming fatigued, thereby allowing the patient to practice walking for longer:

> *I also think logistically, sometimes if you’ve got a patient who requires quite a lot of assistance, particularly with their lower limb, it* [*treadmill training*] *can be much easier because you don’t have to move, you can just sit there while they’re on the treadmill and you can facilitate*. (*Participant 3*)

One physiotherapist spoke about the ability to manipulate the specific aspects of walking when using the treadmill in order to achieve greater therapeutic benefits.

> *I tend to use it for a therapeutic activity when there are very specific things I want. I want to get hip extension, so I will use the treadmill as a means to get that*. (*Participant 14*)

However, some physiotherapists believed that the patterns of muscle activation encouraged by treadmill training are different to those required for overground walking, and therefore believed that treadmill training has the potential to negatively influence walking quality:

> *I mean a lot of that glut and hamstring activation is lost because you’ve got that moving surface underneath*. (*Participant 7*)

In contrast, other therapists felt that there was sufficient research evidence to suggest that mechanically assisted walking training positively influences walking quality:

> *It doesn’t increase spasticity, in fact the evidence is that it actually minimises or reduces spasticity and your primary impairment post-stroke is one of weakness, not increased activity so therefore it negates all of those issues*. (*Participant 14*)

### Practical considerations

Mechanically assisted walking training was described in relation to a number of practical considerations, namely: patient safety, staff efficiency, staff skill in using the treadmill, and access to the necessary equipment. In terms of safety, some therapists endorsed mechanically-assisted walking training as a safer way to mobilize a patient than overground walking; whereas others felt that using a treadmill added an element of unpredictability that could potentially be dangerous:

> *You’ve got to be able to react too, because it’s moving. If you’re facilitating a walk with someone you* [*need to be able to*] *stop if you need to, if it gets unsafe. It’s a lot harder on a treadmill to do that because someone’s got to be hitting the button as well as having hands on. Potentially it needs extra people from a safety perspective*. (*Participant 1*)

While treadmill walking with a harness was identified as one way to improve safety, using this equipment was seen as requiring additional time and assistance to set up. With limited time available for physiotherapy, and limited access to physiotherapy assistants, some physiotherapists perceived that time in therapy was best devoted to other tasks:

> *So if a patient is 3- or 4-assist, then obviously you’ve got those therapists away from the other patients. How much time can you devote to one patient when it’s maybe at the expense of other patients?* (*Participant 10*)

Some physiotherapists cited a lack of skill in training a patient on the treadmill as a potential barrier to treadmill use. Staff in-services, and regularity of practice, fostered confidence to include mechanically assisted walking training as part of a patient’s rehabilitation:

> *A lot of staff don’t know how to use it. No one is teaching it*. [*When I worked in another hospital*] *nobody taught me how to use the treadmill. There was no in-service for the treadmill, there was nothing that happened for the treadmill*. (*Participant 11*)

While mechanically assisted walking training was seen as an effective way to make use of limited gym space, the fact that a facility will often only have access to one treadmill was a barrier to regular treadmill use:

> *We’ve got only one, so that’s a barrier in that way. And there’s a big waiting queue*. (*Participant 11*)

The use of the treadmill appeared to be influenced by patient characteristics, with most therapists stating that not all patients are suitable candidates for mechanically assisted walking training. Therapists thought that heavier people who require more assistance to mobilize, who will require a harness to walk safely, or who need a tilt table to achieve standing may not always be suitable, especially in situations where assistance is limited:

> *People who are more independent in mobility, I think, are better at doing it as well*. (*Participant 1*)
>
> *If you’re in a department where there is fewer staff, it may be much more difficult to use it with some of the lower level patients. A lot of departments would necessarily have to select the patients that are a bit of a higher level because they just physically don’t have the resources to do it*. (*Participant 9*)

Therapists also identified fear as a limiting factor, particularly in older patients who may not have previously used a treadmill:

> *I don’t think a lot of people have been on a treadmill before so there’s that fear as well*. (*Participant 1*)

## DISCUSSION

This study explored the perceptions of physiotherapists surrounding the use of mechanically assisted walking training during the inpatient phase of stroke rehabilitation. The study identified that physiotherapy staff had mixed views of mechanically assisted walking training, and that use is governed by both therapeutic and practical considerations.

Mechanically assisted walking training, as a therapeutic modality, may be underutilized for a number of reasons. First, despite evidence to the contrary (Kuys et al., 2011), some physiotherapists maintained that mechanically assisted walking training encourages maladaptive motor control patterns and may exacerbate impairments, such as spasticity, in people with stroke. Second, a number of staff issues may limit the feasibility of adopting mechanically assisted walking training as part of routine physiotherapy practice. With limited staff available to assist, restricted access to treadmills, and competing demands for time, mechanically assisted walking training may not be seen as a priority. Finally, mechanically assisted walking training is perceived by some as being unsuitable for patients who are heavy and severely disabled and not as an efficient use of time for those who require more assistance. However, mechanically assisted walking training *with body weight* support provided by a harness was originally designed to overcome exactly these factors. By placing the patient in a harness, issues of safety and excessive weight are solved and this method of training is more efficient than overground training (Ada et al 2010) since it results in more walking for the same amount of time.

While we interviewed therapists in Queensland, Australia, other research indicates that our findings may be generalizable to the broader physiotherapy community. McCluskey and colleagues (McCluskey, Vratsistas-Curto, & Schurr, 2013) investigated barriers and enablers to implementing the recommendations of the clinical guidelines by allied health staff at a stroke unit in Sydney, Australia. In particular they focused on practices identified to be priority areas for change, one of which was treadmill training. Barriers to implementing mechanically assisted walking training included concerns for patient’s safety, lack of time and staff, the perception that some patients are not capable of mechanically assisted walking and that other types of training (ie, overground walking) might produce better outcomes. Therapists also reported that the physical demands of assisting patients to walk on the treadmill were a barrier to implementation. Finally, therapists in Sydney conveyed that the absence of prompts to engage in an intervention resulted in failure to adopt it – because mechanically assisted walking training was not offered as part of routine practice, it was often forgotten, despite the fact that physiotherapists were aware of the evidence supporting it (McCluskey et al., 2013). When this study was designed, the Australian clinical stroke guidelines (National Stroke Foundation, 2010) at the time recommended that mechanically assisted walking using a treadmill be implemented to retrain walking and this has not changed in the most recent living guidelines (Stroke Foundation, 2019).

The main limitation of this study is that it was only carried out in two sites. On the other hand, a strength was the breadth of therapist experience included in the focus groups. Mechanically assisted walking training, both with and without body weight support has been included in the Australian clinical guidelines for stroke management since their inception in 2005. It is known that in order to get uptake of clinical guidelines, dissemination of printed materials is not always enough (Giguere et al., 2012). For example, despite the existence of clinical guidelines, in 2018 only 17% of people unable to walk independently after stroke in Australia received mechanically assisted walking training (Stroke Foundation, 2018). Insights gained from the current study can inform strategies for increasing the implementation of mechanically assisted walking training in the future. First, any negative perceptions of mechanically assisted walking training need to be addressed by presentation of evidence of its therapeutic benefit. However, although educational meetings can improve professional practice, it has been suggested that alone they are not likely to be effective for changing complex behaviour (Forsetlund et al., 2009). Given that most of the negative perceptions in the current study were related to practical considerations, it would seem necessary that any educational meeting should include a practical component aimed at upskilling the recipients. Second, audit and feedback may be useful in increasing uptake in clinical practice. It has been suggested that audit and feedback is most effective in situations where baseline performance is low (Ivers et al., 2012), such as is the case with mechanically assisted walking training. Thirdly, the presence of key individuals (such as senior therapists to act as champions) who take responsibility for driving implementation was shown to be important when investigating the uptake of the *The Graded Repetitive Arm Supplementary Program* in the UK (Connell, McMahon, Harris, Watkins, & Eng, 2014). Finally, it is suggested that an integrated framework for translating innovation into practice should include attention to the local, organizational and health system context as well as the characteristics of the recipients and the practicalities of the innovation (Harvey & Kitson, 2016).

In conclusion, therapists’ perceptions of using mechanically assisted walking training during inpatient rehabilitation after stroke were mostly positive in terms of therapeutic consequences but somewhat negative in terms of practical considerations. In order to translate this intervention into clinical practice, a key focus should therefore be on minimizing the perceived practical difficulties.

### Implications for physiotherapy practice

- Mechanically assisted walking training is underutilised in stroke rehabilitation despite evidence of effect
- Physiotherapists had mixed feelings about the therapeutic benefits of mechanically assisted walking training during stroke rehabilitation
- Therapeutic and practical considerations were identified pertaining to using mechanically assisted walking training in stroke rehabilitation and should be addressed in implementation programs

## Data Availability

Data is available on request

## Acknowledgements

This work was supported under the grant: Health and Medical Research Physiotherapy Project Grant, Queensland Health, Australia (ID QCOS/018820). We would like to thank all staff who voluntarily shared their views, and Dr Robyn Lamont for conducting the focus groups.

